# Evaluation of attitude among infertile couples about continuing assisted reproductive technologies therapy during novel coronavirus outbreak

**DOI:** 10.1101/2020.09.01.20186320

**Authors:** Sepideh Peivandi, Alireza Razavi, Shervin Shafiei, Marzieh Zamaniyan, Asma Orafaie, Hamed Jafarpour

## Abstract

**Study question:** Does the fear of the coronavirus disease 2019 (COVID-19) pandemic reduce the desire of infertile couples to continue treatment?

**Summary answer:** Most of the participants in this study wanted to continue treatment.

**What is known already:** The effect of the prevalence of infectious diseases including the Zika virus on the attitude of infertile couples has been studied in very few studies. However, the effect of the outbreak of COVID-19 on the attitude of infertile couples has not been investigated.

**Study design, size, duration:** We conducted a prospective longitudinal study on forty-six infertile couples (n=92) who referred to our infertility clinic from 4 March 2020 through 20 June 2020.

**Participants/materials, settings, methods:** This study is based on potential infertile couples for treatment with assisted reproductive technology (ART) who referred to our infertility clinic, Sari, Iran (median age of 35.5 years). All individuals with primary infertility, as defined by the World Health Organization (WHO) and candidates for ART, were included in the study. People who did not agree to participate in the study were excluded. Subjects were surveyed using a researcher-made questionnaire. This questionnaire has four sections as follows: The first part included demographic information and clinical characteristics, the second part included medical records, the third part included questions related to assessing the level of awareness regarding coronavirus infection, and the fourth part included questions related to the attitude towards continuing infertility treatment. The validity of this questionnaire was assessed by three infertility specialists and was confirmed with Cronbach’s alpha of 0.78. A P-value of less than 0.05 was considered statistically significant.

**Main results and the role of chance:** There is no significant relationship between COVID-19 symptoms and the level of awareness (P-value <0.05). Thirty-two patients (33.33%) had decreased motivation to continue treatment during COVID-19 pandemic. Fear of transmission to the fetus (28.13%) had the highest frequency among the causes of decreased motivation to continue treatment (P-value = 0.011). Confidence in support of the treatment team (56.67%) was the most common reason for lack of motivation in people without decreased motivation (P-value <0.001). The majority of patients had a very high or high tendency (65.22%) to continue or start treatment during the COVID-19 pandemic (P-value <0.001). Most people had an average fear of getting the disease (39.13%) (value <0.001). Examining the relationship between the presence of COVID-19 symptoms and attitude level variables showed that there is only a significant relationship between the greater desire to have a child and the continuation or initiation of treatment with the presence of COVID-19 symptoms (P-value = 0.032).

**Limitations, reasons for caution:** We were not able to fully assess patients’ deep feelings and cultural beliefs, due to the use of questionnaires and the lack of interviews.

**Wider implications of the findings:** Our results showed for the first time that patients’ knowledge about COVID-19 and trust in the treatment staff played an effective role in selecting and continuing infertility treatment. To clarify this issue, studies with the larger statistical community in the form of interviews are needed.

**Study funding/completing interest(s):** The study received financial support from the Mazandaran University of Medical Sciences (Grant number: 7903). None of the funders had any role in the study design, collection, analysis, or interpretation of data, writing of the paper, or publication decisions. The authors have nothing to declare.

**Trial registration number:** N/A

## Introduction

In December 2019, unknown pneumonia was reported in Wuhan, China, which belongs to the coronavirus family (1, 2). The most important way of severe acute respiratory syndrome coronavirus 2 (SARS-CoV-2) transmission is through direct person-to-person contact and respiratory droplets (3). In addition, this virus was also detected in urine, feces, and tears, which confirm the possibility of transmission through the fecal-oral route (4, 5). So far, there is no evidence that this disease can be transmitted through sexual secretions and the pregnant mother to the child, but there are doubts and suspicions in this regard (6, 7).

Bioinformatics analysis has shown that the angiotensin-converting enzyme 2 (ACE2), the receptor used by SARS-CoV-2 for infectivity, is abundantly expressed in the testes and male reproductive system (8, 9). There are also reports that Human coronavirus 229E (HCoV-229E), which is a member of the SARS-CoV-2 family, has been found in vaginal discharge (10). Therefore, it is suspected that SARS-CoV-2 could be transmitted through sexual secretions (7), and such concerns about infectious diseases can affect the process of continuing infertility treatment in infertile couples (11, 12). Infertility is a couple’s failure to conceive after at least one year of trying (13). Worldwide, the rate of primary infertility in women aged 20-44 is estimated at 1.9%. However, the prevalence of infertility varies from country to country (14).

According to data collected by the World Health Organization (WHO) in 2004, there are 187 million infertile couples in developing countries except for China. However, a large part of the world, including Iran, is reluctant to use modern methods of pregnancy (15, 16). On average, about 10-15% of couples in Iran have infertility problems (17), of which 50% seek medical treatment, which may initially include medication or surgery (18). If these first treatments do not work or are considered inappropriate, about 3% of these couples are recommended to use assisted reproductive technologies (ART). One of these methods is *in vitro* fertilization (IVF), which accounts for more than 99% of ARTs. The success rate of this method is 16.6-20.2% (19). When starting treatment, couples have to endure a variety of treatments, including ovarian stimulation, regular monitoring, egg retrieval, embryo transfer, and the use of progesterone supplements (20). Examination, research, and injections can severely disrupt a couple’s daily routine (21). Therefore, couples can be expected to suffer from the stressful experience of infertility because they endure the pressure of painful treatment and the fear of failure (12). It has also been found that couples seeking IVF approach are more likely than non-IVF couples to report unstable relationship due to length and treatment expectations (22, 23). While the social, ethical, and legal issues regarding ART are well documented globally, as well as in Asia, many Iranians are unaware of the exact application of this technology (16). Due to the nature of treatment and pregnancy, objective measurement of psychological, social, emotional, and attitudinal effects on couples using ART is limited (12).

Several studies have examined the experiences of infertile couples in IVF treatment (23). These studies have mainly focused on the impact of social contexts on infertility, especially in the two weeks waiting for pregnancy outcomes following embryo transfer (24) and life after unsuccessful treatment (25). Also, it has been reported that some viral infectious diseases such as Zika can cause abnormalities and microcephaly in the fetus (26), which has influenced the attitude and decision of infertile couples to continue infertility treatment with ART (26, 27).

Due to the fact that the transmission of SARS-CoV-2 through sexual secretions, and through the pregnant mother to the child has not yet been confirmed, during coronavirus disease 2019 (COVID-19) pandemic, some of these important aspects need to be considered: 1) Accurate identification Infertile women who are sensitive at this time; 2) effective personalization of stimulation based on maternal age and ovarian reserve; and 3) prevention of ART-related risks such as ovarian hyperstimulation syndrome (OHSS), complications associated with egg retrieval, and multiple pregnancies (28).

All of these factors influence couples’ decisions to seek infertility treatment. Considering that no study has been done on the effect of COVID-19 pandemic on the decision of infertile couples to continue infertility treatment and the importance of investigating this issue in Iran, we decided to examine the attitude and knowledge of such individuals regarding the treatment during COVID19 pandemic.

## Methods

### Study design and participants

This study is a prospective longitudinal investigation on potential infertile couples for treatment with ARTs who referred to our infertility clinic from 4 March 2020 to 20 June 2020. Forty-six infertile couples (n=92) were studied voluntarily and anonymously in this study. This study was performed after the approval of the Biomedical Research Ethics Committee of Mazandaran University of Medical Sciences (code: IR.MAZUMS.REC.1399.03).

All individuals with primary infertility, as defined by the WHO (29) and candidates for assisted reproductive technology (ART), were included in the study. People who did not agree to participate in the study were excluded. Subjects were surveyed using a researcher-made questionnaire. This questionnaire has four sections as follows: The first part included demographic information and clinical characteristics, the second part included medical records, the third part included questions related to assessing the level of awareness regarding coronavirus infection, and the fourth part included questions related to the attitude towards continuing infertility treatment. The validity of this questionnaire was assessed by three infertility specialists and was confirmed with Cronbach’s alpha of 0.78.

### Statistical analysis

Quantitative data were analyzed using Kolmogorov-Smirnov and Shapiro-Wilk tests, *t*-test, and Pearson correlation coefficient. Data were also analyzed through the Q–Q (quantile-quantile) plot, as well as skewness and elongation index. A P-value of less than 0.05 was considered statistically significant.

## Results

A total of 92 patients (46 couples) were enrolled. The mean age was 35.51±5.51 years with a minimum age of 22 and a maximum age of 50 years. The mean age of men (37.35±5.24) was significantly higher than women (33.67 ± 5.18) (P-value = 0.001). The demographic characteristics of the patients are shown in Table 1.

**Table 1.** Frequency of demographic characteristics

| Variables |  | P-value |
| --- | --- | --- |
| **Age, Mean (SD), years** |  |  |
| Man | 37.35 (5.24) | 0.001 |
| Women | 33.67 (5.18) |  |
| **Level of Education, N (%)** |  |  |
| Illiterate | 2 (2.17) | <0.001 |
| Less than diploma and diploma | 28 (30.43) |  |
| Bachelor | 45 (48.91) |  |
| Masters up | 17 (18.48) |  |
| **Living area, N (%)** |  |  |
| City | 72 (78.26) | <0.001 |
| Village | 20 (21.74) |  |
| **Occupation, N (%)** |  |  |
| Unemployed | 29 (31.52) | 0.001 |
| University student | 12 (13.04) |  |
| With a fixed income | 32 (34.78) |  |
| No fixed income | 19 (20.65) |  |
| **Monthly income, N (%), Million rials** |  |  |
| Less than 20 | 35 (38.04) | <0.001 |
| 20 | 24 (26.09) |  |
| 20 to 50 | 24 (26.09) |  |
| More than 5 | 9 (9.78) |  |
| **Religion, N (%)** |  |  |
| Muslim-Shia | 89 (96.74) | <0.001 |
| Muslim-Sunni | 3 (3.26) |  |
| **History of underlying disease, N (%)** | 6 (6.52) | <0.001 |

Most of women did not have a previous pregnancy, delivery, and abortion (P-value <0.001). Four women (4.35%) had a history of multiple births. None of the people had a history of stillbirth. Four and 11 couples had a history of successful and unsuccessful infertility treatment, respectively. Fifteen couples with a history of infertility treatment. The treatment methods used by 15 couples with previous treatment history included IUI (66.67%), embryo transfer (86.66%), follicle aspiration (40.00%) (P-value <0.001). Only two patients (2.17%) were experiencing symptoms of COVID-19 and no one was hospitalized.

Table 2 shows the frequency of answers to the questionnaire of the level of awareness of the patients. The highest frequency of information acquisition method was radio and television (43.48%) (P-value <0.001). The majority of people chose to use a mask (55.43%) as the most effective way of prevention (P-value <0.001). There is no significant relationship between COVID-19 symptoms and the level of awareness (P-value <0.05).

**Table 2.**
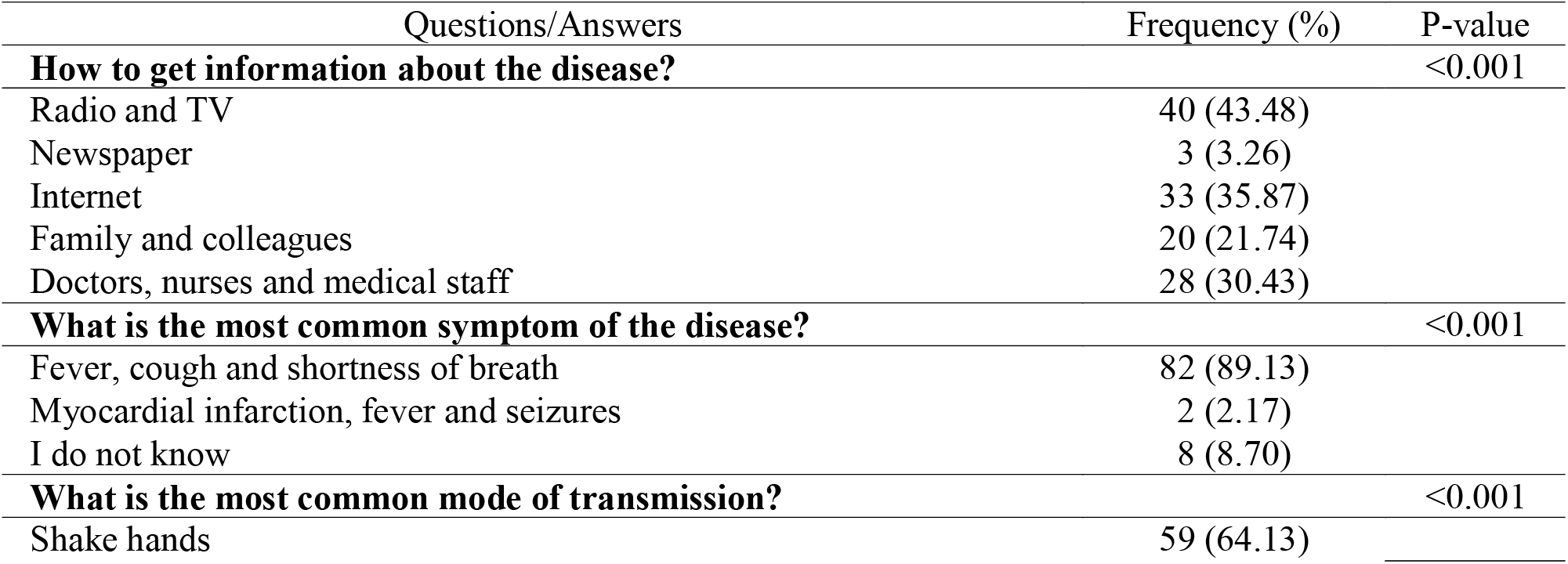
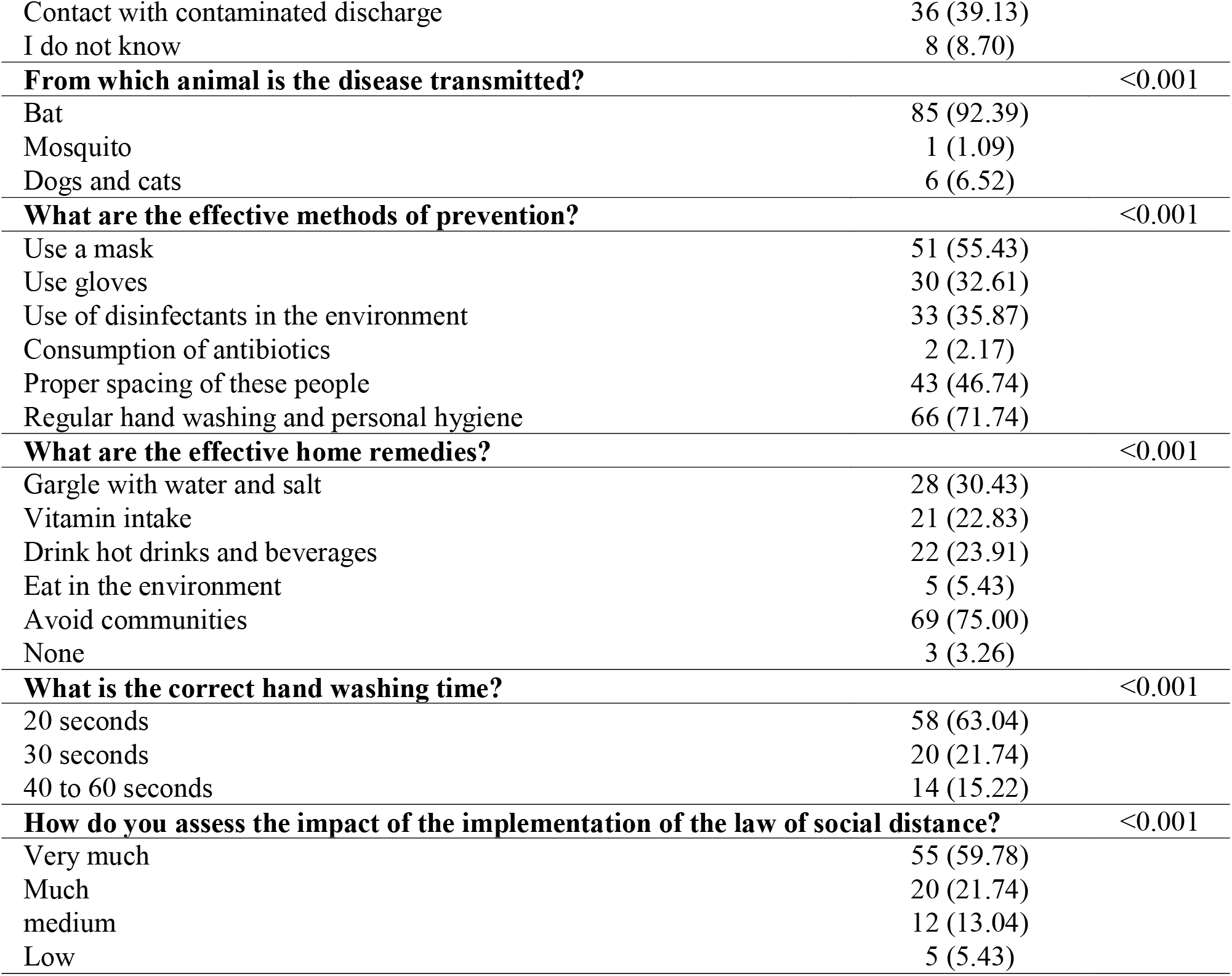
Frequency of answers to questions of the level of awareness

Thirty-two patients (33.33%) had decreased motivation to continue treatment during COVID-19 pandemic. Fear of transmission to the fetus (28.13%) had the highest frequency among the causes of decreased motivation to continue treatment (P-value = 0.011). Confidence in support of the treatment team (56.67%) was the most common reason for lack of motivation in people without decreased motivation (P-value <0.001) (Table 3).

**Table 3.** Frequency (%) of causes of decreased motivation to continue infertility treatment in patients during the COVID-19 pandemic

| Decreased motivation | Causes | Frequency (%) | P-value |
| --- | --- | --- | --- |
| Yes<br>(n=32) | Fear of transmitting the disease to the fetus | 9 (28.13) | 0.011 |
|  | Fear of getting sick during treatment | 7 (21.88) |  |
|  | Fear of the negative effect of the virus on the outcome of pregnancy | 4 (12.50) |  |
|  | Fear of unknown effects of the virus in pregnancy | 5 (15.63) |  |
|  | Fear of transmitting the disease | 7 (21.88) |  |
| No<br>(n=60) | Desire to have children even if there is a risk of transmission of infection | 10 (16.67) | <0.001 |
|  | Time limit for treatment | 16 (26.67) |  |
|  | Trust the support of the treatment team | 34 (56.67) |  |

The majority of patients had a very high or high tendency (65.22%) to continue or start treatment during the COVID-19 pandemic (P-value <0.001). Most people had an average fear of getting the disease (39.13%) (value <0.001). Fear of getting disease due to presence in the treatment environment in most people (39.13%) was moderate (value <0.001). Fear of the negative impact of the coronavirus on pregnancy (35.87%) was also moderate in most people (value <0.001) (Figure 2). Examining the relationship between the presence of COVID-19 symptoms and attitude level variables showed that there is only a significant relationship between the greater desire to have a child and the continuation or initiation of treatment with the presence of COVID-19 symptoms (P-value = 0.032).

**Figure 1.**
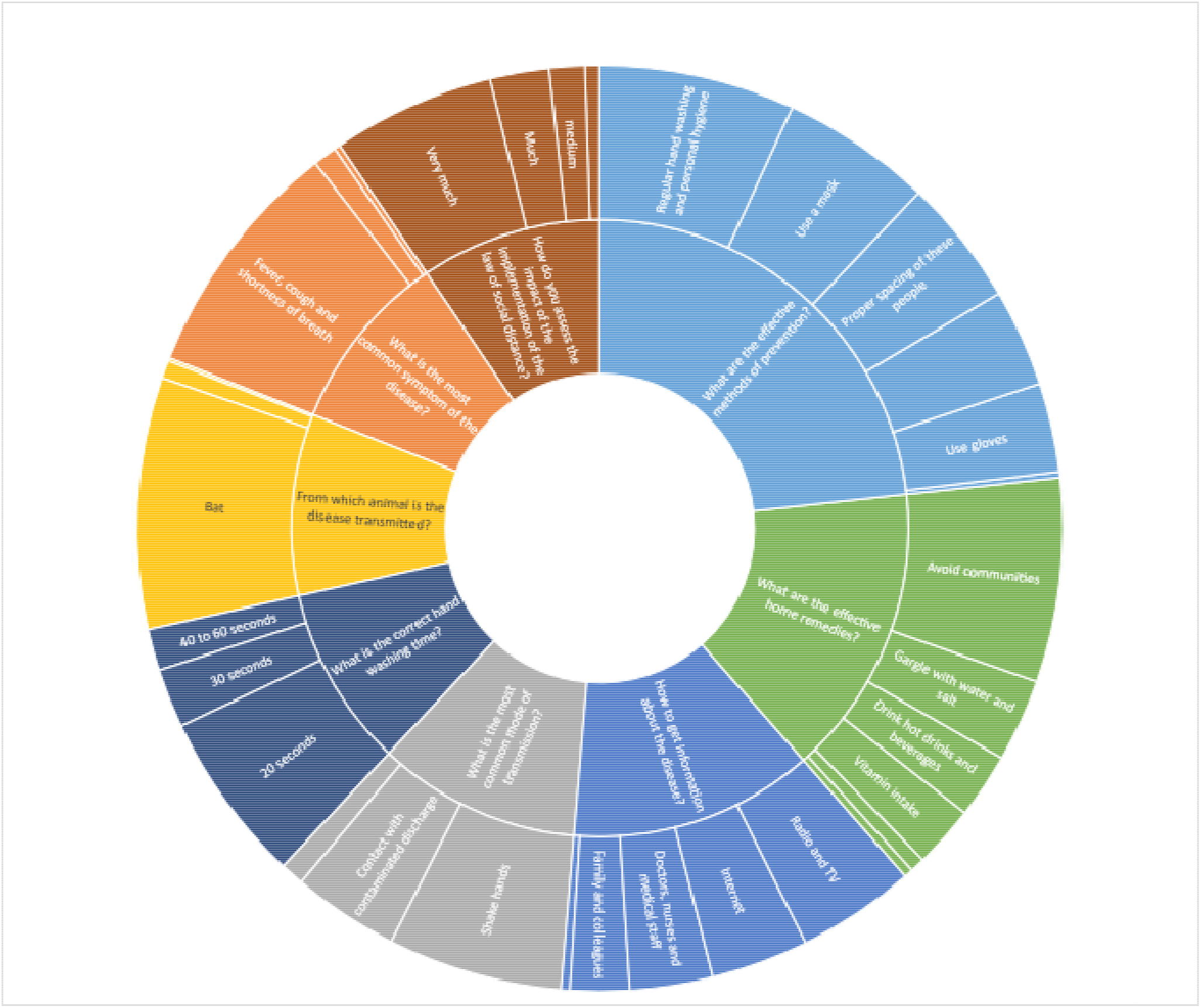
Sunburst chart of answers to questions of level of awareness

**Figure 2.**
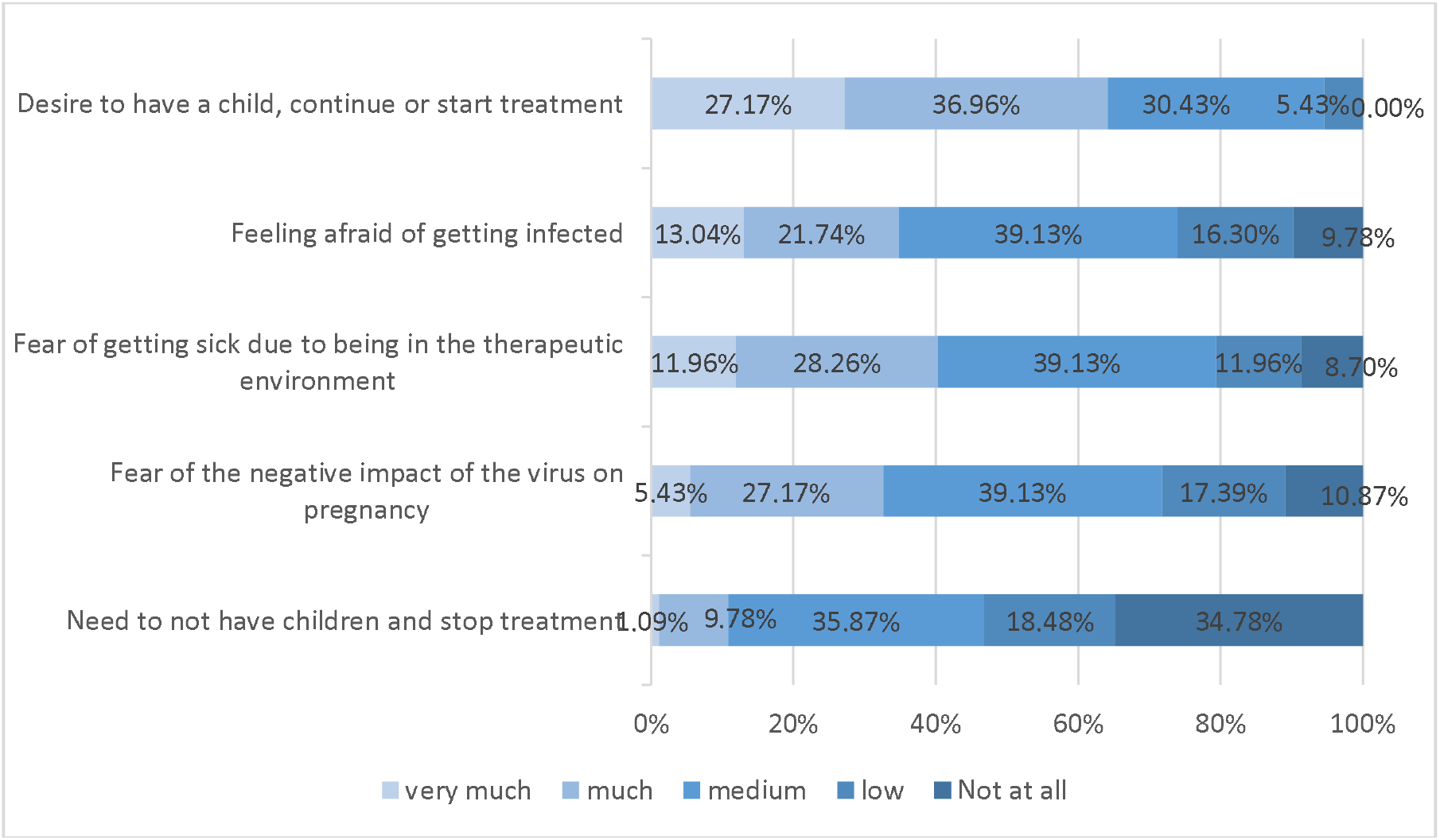
Frequency of attitude level factors of coronavirus infection

## Discussion

To our knowledge, this cross sectional study is the first to investigate the attitudes and consciousness of infertile couples, candidate therapy about continuing treatment during COVID19 outbreak. The level of knowledge and awareness of infertile couples and their ability to fully respond to COVID-19 in their communities is somewhat limited (29). In this study, among 92 patients (46 couples), we found that more than 60% of individuals did not have a reduced motivation to continue treatment. One-third of patients who had a decreased motivation to continue treatment felt the need because of their infection and the transmission of the disease to the fetus and others. Also, in the present study, there was no significant relationship between the presence of COVID-19 symptoms and the level of awareness of couples. There was only a significant association between COVID-19 symptoms and a greater desire to have children and to continue or start treatment. In our patients, subjects with fixed salaries, bachelor’s degrees, and Shiite religion had the highest frequency. Also, the number of individuals who had no history of underlying diseases and lived in the city was significantly higher. In our survey, sociocultural status had an effect on couples’ attitudes. These results are reinforced by some studies (30). Since infertility treatment is very expensive and some couples need years of treatment and also some of them have to travel to another city which is more expensive for treatment, these issues can affect their attitude and it can be claimed that the socio-economic situation may change the couple’s attitude (16). But in this study, the majority of clients had an income less than 100$; so, there was not a great difference between their incomes, and there was not a statistical difference between their attitudes.

In our study, most people were nulliparas and had no history of infertility treatment. Therefore, in this study, no relationship can be imagined between pregnancy history and infertility treatment with patients’ attitudes toward continuing treatment. Most people wanted to use intrauterine insemination (IUI) and embryo transfer for their treatment. IUI and embryo transfer are relatively simple, common, and safe procedures, and the risk of serious complications is low (31, 32). Although some studies have shown that the process of treating infertility, including ART and related tests, is very long and tedious (33, 34), in our study most patients, despite the fear of the COVID-19 pandemic, were tended to perform and continue their treatment.

In our study, about 24% of patients experienced unsuccessful infertility treatment. In some studies, unsuccessful infertility treatment not only led to immediate heart failure and shock, but also to long-term psychological trauma. In these surveys, pregnant women reported experiencing severe anxiety due to previous unsuccessful IVF attempts (35, 36). In addition, we found that more than half of those who wished to continue infertility treatment trusted the support of the treatment team. It seems that trust in the treatment team and the performance of the treatment team members play an important role in increasing the desire of infertile couples to continue infertility treatment. This factor, ie the support of the treatment team for patients, has been studied in some studies. For example, the lack of counseling time in the clinic and the lack of psychological support have been assessed as insufficient in health care providers. In fact, the busy schedule of a clinic makes it difficult for the clinic to provide information and psychological needs to outpatients (37). Research has shown that the support of health professionals is very effective in improving the stress and anxiety of people with infertility (38).

In our study, couples’ knowledge, and awareness of general information about COVID-19 such as common symptoms, prevention methods, and also about whether SARS-CoV-2 is transmitted vertically or sexually (0.00%), was evaluated well. Many people believed that social distance could help curb this pandemic. The response of many patients was influenced by the information published in the media, so that the role of the media in this result was very significant. In our patients, due to the knowledge of most people in this study that the virus is not transmitted vertically, the majority of people were very, very willing to continue or start treatment during the pandemic of this disease. The highest prevalence of fear among people whose motivation to continue treatment was reduced was due to fear of developing COVID-19 due to being in the treatment environment and feeling afraid of the negative impact of the virus on pregnancy. A noteworthy point in our study was that there was only a significant association between COVID19 symptoms and a greater desire to have children and to continue or start treatment. Some studies have shown that the knowledge gap among infertile women in the pandemic of infectious diseases causes a difference in their attitudes in choosing the type of treatment and its continuation (39).

## Limitations

The sample size of our study was small. Although data saturation has been reached, future research with larger sample sizes is needed to address other factors that may affect the experience of infertile couples and reach well-appointed conclusions. Another limitation was the use of a questionnaire, which may prevent the deep expression of deep feelings. Using interviews to gather information from a couple in person will allow researchers to examine the dual relationships between couples. Studying a more homogeneous group allows researchers to gain a deeper understanding of the cultural context and its effects on infertile couples’ attitudes toward continuing treatment. Studies comparing the differences between infertile couples in different cultural groups in Iran can be helpful in the future. Given this, the results of this study cannot be generalized to all people who use ART or in all regions of Iran, although it may be applicable in similar fields.

## Conclusion

In our study, most patients had acceptable knowledge about COVID-19 and how it was transmitted. Due to the knowledge of most people in this study that the virus is not transmitted vertically, most people in the study wanted to continue their infertility treatment in the COVID19 pandemic.

### Supplementary data

Supplementary data are available at Human Reproduction online.

## Data Availability

All data is available in the given manuscript.

## Acknowledgment

Special thanks to Sexual and Reproductive Health Research Center of Imam Khomeini Hospital and Student Research Committee of Mazandaran University of Medical Sciences to supporting to project.

## Authors’ roles

S.P conceived the study design, completed the data analysis, and wrote the article. A.R significantly contributed to the conception of the study design and interpretation of the data, critically revised the article, and approved the final version. H.J significantly contributed to the conception of the study design and analysis, critically revised the article, and approved the final version. S.S significantly contributed to the conception of the study design and analysis, critically revised the article, and approved the final version. M.Z significantly contributed to the conception of the study design and interpretation of the data, critically revised the article, and approved the final version. A.O significantly contributed to the conception of the study design and interpretation of the data, critically revised the article, and approved the final version.

## References

1. Davoodi L, Jafarpour H, Taghavi M, Razavi A. COVID-19 Presented With Deep Vein Thrombosis: An Unusual Presenting. Journal of Investigative Medicine High Impact Case Reports. 2020;8:2324709620931239.

2. Davoodi L, Jafarpour H, Kazeminejad A, Soleymani E, Akbari Z, Razavi A. Hydroxychloroquine-induced Stevens–Johnson syndrome in COVID-19: a rare case report. Oxford Medical Case Reports. 2020;2020(6).

3. Razavi A, Davoodi L, Shojaei L, Jafarpour H. COVID-19 in Children: A Narrative Review. Open Access Macedonian Journal of Medical Sciences. 2020;8(T1):23-31.

4. Guan W-j, Ni Z-y, Hu Y, Liang W-h, Ou C-q, He J-x, et al. Clinical characteristics of coronavirus disease 2019 in China. New England journal of medicine. 2020;382(18):1708–20.

5. Peng L, Liu J, Xu W, Luo Q, Chen D, Lei Z, et al. SARS-CoV-2 can be detected in urine, blood, anal swabs, and oropharyngeal swabs specimens. Journal of Medical Virology. 2020;92(9):1676–80.

6. Song C, Wang Y, Li W, Hu B, Chen G, Xia P, et al. Absence of 2019 novel coronavirus in semen and testes of COVID-19 patients†. Biology of Reproduction. 2020;103(1):4–6.

7. Patrì A, Gallo L, Guarino M, Fabbrocini G. Sexual transmission of severe acute respiratory syndrome coronavirus 2 (SARS-CoV-2): A new possible route of infection? J Am Acad Dermatol. 2020;82(6):e227.

8. Wang Z, Xu X. scRNA-seq Profiling of Human Testes Reveals the Presence of the ACE2 Receptor, A Target for SARS-CoV-2 Infection in Spermatogonia, Leydig and Sertoli Cells. Cells. 2020;9(4):920.

9. Chen F, Lou D. Rising Concern on Damaged Testis of COVID-19 Patients. Urology. 2020;142:42.

10. Gagneur A, Dirson E, Audebert S, Vallet S, Legrand-Quillien M, Laurent Y, et al. Materno-fetal transmission of human coronaviruses: a prospective pilot study. European Journal of Clinical Microbiology & Infectious Diseases. 2008;27(9):863–6.

11. Semprini AE, Hollander LH, Vucetich A, Gilling-Smith C. Infertility treatment for HIV-positive women. Women’s health. 2008;4(4):369–82.

12. El Kissi Y, Romdhane AB, Hidar S, Bannour S, Idrissi KA, Khairi H, et al. General psychopathology, anxiety, depression and self-esteem in couples undergoing infertility treatment: a comparative study between men and women. European Journal of Obstetrics & Gynecology and Reproductive Biology. 2013;167(2):185–9.

13. Mousavi SA, Masoumi SZ, Keramat A, Pooralajal J, Shobeiri F. Assessment of questionnaires measuring quality of life in infertile couples: a systematic review. Journal of reproduction & infertility. 2013;14(3):110.

14. Mascarenhas MN, Flaxman SR, Boerma T, Vanderpoel S, Stevens GA. National, regional, and global trends in infertility prevalence since 1990: a systematic analysis of 277 health surveys. PLoS Med. 2012;9(12):e1001356.

15. Kian EM, Riazi H, Bashirian S. Attitudes of Iranian infertile couples toward surrogacy. Journal of human reproductive sciences. 2014;7(1):47.

16. Afshani SA, Abdoli AM, Hashempour M, Baghbeheshti M, Zolfaghari M. The attitudes of infertile couples towards assisted reproductive techniques in Yazd, Iran: A cross sectional study in 2014. International Journal of Reproductive BioMedicine. 2016;14(12):761.

17. Bokaie M, Farajkhoda T, Enjezab B, Heidari P, Zarchi MK. Barriers of child adoption in infertile couples: Iranian’s views. Iranian journal of reproductive medicine. 2012;10(5):429.

18. Greil AL, Slauson-Blevins K, McQuillan J. The experience of infertility: a review of recent literature. Sociology of health & illness. 2010;32(1):140–62.

19. Sullivan EA, Zegers-Hochschild F, Mansour R, Ishihara O, De Mouzon J, Nygren K, et al. International Committee for Monitoring Assisted Reproductive Technologies (ICMART) world report: assisted reproductive technology 2004. Human Reproduction. 2013;28(5):1375–90.

20. Casanova R. Beckmann and Ling’s Obstetrics and Gynecology: Lippincott Williams & Wilkins; 2018.

21. Pasch LA, Christensen A. Couples facing fertility problems. The psychology of couples and illness: Theory, research, & practice. Washington, DC, US: American Psychological Association; 2000. p. 241-67.

22. Burns LH. Parenting after infertility. Infertility counseling: A comprehensive handbook for clinicians, 2nd ed. New York, NY, US: Cambridge University Press; 2006. p. 459-76.

23. Wang K, Li J, Zhang JX, Zhang L, Yu J, Jiang P. Psychological characteristics and marital quality of infertile women registered for in vitro fertilization-intracytoplasmic sperm injection in China. Fertility and sterility. 2007;87(4):792–8.

24. Lampley TM. The interim window: Women’s experiences during in vitro fertilization leading to maternal embryo attachment: University of Nevada, Las Vegas; 2010.

25. Su T, Chen Y. Transforming hope: the lived experience of infertile women who terminated treatment after in vitro fertilization failure. Journal of nursing Research. 2006;14(1):46.

26. Vaiarelli A, Bulletti C, Cimadomo D, Borini A, Alviggi C, Ajossa S, et al. COVID-19 and ART: the view of the Italian Society of Fertility and Sterility and Reproductive Medicine. Reproductive BioMedicine Online. 2020;40(6):755–9.

27. Ying LY, Wu LH, Loke AY. The Experience of Chinese Couples Undergoing In Vitro Fertilization Treatment: Perception of the Treatment Process and Partner Support. PLoS One. 2015;10(10):e0139691.

28. Sohrabvand F, Jafarabadi M. Knowledge and attitudes of infertile couples about assisted reproductive technology. International Journal of Reproductive BioMedicine. 2005;3(2):90–0.

29. Vaiarelli A, Bulletti C, Cimadomo D, Borini A, Alviggi C, Ajossa S, et al. COVID-19 and ART: the view of the Italian Society of Fertility and Sterility and Reproductive Medicine. Reproductive BioMedicine Online. 2020.

30. Suzuki K, Hoshi K, Minai J, Yanaihara T, Takeda Y, Yamagata Z. Analysis of national representative opinion surveys concerning gestational surrogacy in Japan. European Journal of Obstetrics & Gynecology and Reproductive Biology. 2006;126(1):39–47.

31. Scotland GS, McNamee P, Peddie V, Bhattacharya S. Safety versus success in elective single embryo transfer: women’s preferences for outcomes of in vitro fertilisation. BJOG: An International Journal of Obstetrics & Gynaecology. 2007;114(8):977–83.

32. Lenton E. Stimulated intrauterine insemination: efficient, cost-effective, safe? Human Fertility. 2004;7(4):253–65.

33. Cipolletta S, Faccio E. Time experience during the assisted reproductive journey: A phenomenological analysis of Italian couples’ narratives. Journal of Reproductive and Infant Psychology. 2013;31(3):285–98.

34. Bolvin J, Lancastle D. Medical waiting periods: imminence, emotions and coping. Women’s Health. 2010;6(1):59–69.

35. Volgsten H, Svanberg AS, Olsson P. Unresolved grief in women and men in Sweden three years after undergoing unsuccessful in vitro fertilization treatment. Acta obstetricia et gynecologica Scandinavica. 2010;89(10):1290–7.

36. Ying L-Y, Wu LH, Loke AY. The experience of Chinese couples undergoing in vitro fertilization treatment: perception of the treatment process and partner support. PloS one. 2015;10(10):e0139691.

37. Widge A. Seeking conception: Experiences of urban Indian women with in vitro fertilisation. Patient Education and Counseling. 2005;59(3):226–33.

38. Brucker PS, McKenry PC. Support from health care providers and the psychological adjustment of individuals experiencing infertility. Journal of Obstetric, Gynecologic, & Neonatal Nursing. 2004;33(5):597–603.

39. Dickson DA, Mankee-Sookram S, Jess N, Minto-Bain CL, Ramsewak S. Knowledge, attitudes and practices of patients of a fertility clinic in a ZIKA-endemic Caribbean country. Fertility and Sterility. 2017;108:e327-e8.

